# Validation of the Strengths and Difficulties Questionnaire (SDQ) emotional subscale in assessing depression and anxiety across development

**DOI:** 10.1101/2023.03.30.23287961

**Authors:** Jessica M Armitage, Foteini Tseliou, Lucy Riglin, Charlotte Dennison, Olga Eyre, Rhys Bevan Jones, Frances Rice, Ajay K Thapar, Anita Thapar, Stephan Collishaw

**Affiliations:** Wolfson Centre for Young People’s Mental Health, Cardiff University, Wales, United Kingdom; Division of Psychological Medicine and Clinical Neurosciences, Centre for Neuropsychiatric Genetics and Genomics, Cardiff University, Wales, United Kingdom

**Keywords:** SDQ, emotional problems, depression, anxiety, measurement, ALSPAC

## Abstract

Emotional disorders are common in childhood, and their prevalence sharply increases during adolescence. The Strengths and Difficulties Questionnaire (SDQ) is widely used for screening emotional and behavioural difficulties in children and young people, but little is known about the accuracy of the emotional subscale (SDQ-E) in detecting emotional disorders, and whether this changes over development. Such knowledge is important in determining whether symptom changes across age are due to developmental or measurement differences. This study assessed the validity of the SDQ-E and two individual items (low mood and general worry) in differentiating between cases and non-cases of Major Depressive Disorder (MDD), Generalised Anxiety Disorder (GAD), and other anxiety disorders across ages 7, 10, 13, 15, and 25 years in a UK population cohort. Analyses showed moderate accuracy of the subscale in discriminating cases of MDD (AUC=0.67-0.85), and high accuracy for discriminating cases of GAD (AUC=0.80-0.93) and any anxiety disorder (AUC=0.74-0.83) compared to non-cases. The SDQ-E performed well across ages and sex, and generally performed better than the two individual items. Together our findings validate the SDQ-E as a screen for emotional disorders during childhood, adolescence, and early adulthood, and as a tool for longitudinal research on depression and anxiety disorders.

## 1. Introduction

Emotional disorders like depression and anxiety are common in childhood, and their prevalence sharply rises in adolescence and early adult life (Thapar et al., 2022). In order to understand how emotional problems develop in young people, it is crucial that symptoms are assessed repeatedly using comparable measures. Yet a current barrier to longitudinal research on the developmental course of emotional disorders is that different measures of depression and anxiety are typically used for adolescents and adults. This makes it difficult to determine whether any differences are developmental or due to measurement changes.

The Strengths and Difficulties Questionnaire (SDQ) is the most widely used screen for emotional and behavioural difficulties in children and young people (Goodman, 2001). It includes five subscales (emotional symptoms, hyperactivity/inattention, conduct and peer relationship problems, prosocial behaviour) that can be either combined into a total difficulties score, or investigated as separate subscales. The SDQ emotional subscale (SDQ-E) is made up of five items relating to depression, worry, fear, nervousness, and somatic symptoms. The measure has previously been validated against diagnostic measures of depression or anxiety disorders (e.g. Goodman et al., 2003), but it is unclear whether the SDQ-E (or its constituent items) show different patterns of association with depression and anxiety at different developmental time points.

Much of the research on the validity of the SDQ in identifying depression or anxiety disorders has focused on either the full SDQ scale (Kuhn et al., 2017), the emotional subscale in childhood alone (Silva et al., 2015), or has grouped children and adolescents into one age category (Goodman et al., 2001; Goodman et al., 2003). While such research has provided initial promising evidence for the SDQ-E in detecting emotional disorders, we know little about the validity of the SDQ-E in detecting Major Depressive Disorder (MDD) and anxiety disorders across different ages. Such knowledge is important for longitudinal research that aims to retain the same measure to assess the developmental course of depression and anxiety. Understanding whether one scale, or a reduced set of items, present similar levels of accuracy across development could also help confirm existing research on changes in incidence, which show that MDD and some anxiety disorders have a marked increase in prevalence across adolescence, particularly for females (Kessler et al., 2005; Thapar et al., 2022). If measurement scales used to assess these disorders are valid across development, we can be confident that symptom changes are developmental and not due to measurement changes.

The aims of the current study were to assess and compare the validity of the emotional subscale and individual items that reflect core features of depression or anxiety (low mood and general worry respectively) for detecting diagnoses of DSM-5 (American Psychiatric Association, 2013) MDD, Generalised Anxiety Disorder (GAD), as well as other anxiety disorders across development. To do this we use a UK population cohort that includes participants who have been assessed repeatedly using the SDQ, and have completed research diagnostic instruments designed to generate clinical diagnoses of depression and anxiety. The study represents the first of its kind to test (a) the discriminative validity of the emotional subscale in detecting depressive and anxiety disorders across development, and (b) whether individual depressive or worry items from the emotional subscale are able to differentiate those with depressive or anxiety disorders from those without, at levels similar to those for the full SDQ emotional subscale. Doing so could aid future developmental research in selecting appropriate measures across developmental periods. This could prove especially important for improving the assessment and monitoring of these disorders across development and during the transition from child and adolescent to adult health services.

## 2. Methods

### 2.1. Sample

Data were taken from the Avon Longitudinal Study of Parents and Children (ALSPAC), a prospective, longitudinal birth cohort study based in the UK (Boyd et al., 2013; Fraser et al., 2013). Pregnant women based in the Avon area of England, with an expected delivery date between 1st April 1991 and 31st December 1992, were eligible for the study (Fraser et al., 2013). Of these, 20,248 pregnancies were identified as being eligible and the initial number of pregnancies enrolled was 14,541, with 13,988 children alive at 1 year of age. The sample for analyses using variables from age 7 and onwards include 15,447 pregnancies following further follow-up (Northstone et al., 2019). Of these, 14,901 children were alive at 1 year of age. Study data from 22 years were collected and managed using REDCap electronic data capture tools hosted at the University of Bristol (Harris et al., 2009). REDCap (Research Electronic Data Capture) is a secure, web-based software platform designed to support data capture for research studies. Please note that the study website contains details of all the data that is available through a fully searchable data dictionary and variable search tool: http://www.bristol.ac.uk/alspac/researchers/our-data/.

Ethical approval for the study was obtained from the ALSPAC Ethics and Law Committee and the Local Research Ethics Committees. Informed consent for the use of data collected via questionnaires and clinics was obtained from participants following the recommendations of the ALSPAC Ethics and Law Committee at the time. Details of the samples included can be found in Supplementary Tables 1 and 2. Where families included multiple births, we only included the oldest sibling, as per previous research on this sample (Eyre et al., 2021).

### 2.2. Measures

The Strengths and Difficulties Questionnaire emotional subscale (SDQ-E) and the Development and Wellbeing Assessment (DAWBA) were administered on five occasions: at approximate ages 7 years (mean ages: 81 and 91 months respectively), 10 years (115 and 128 months), 13 years (157 months and 166 months), 15/16 years (198 and 185 months), and at 25 years.

#### 2.2.1. Strengths and Difficulties Questionnaire (SDQ)

The Strengths and Difficulties Questionnaire (SDQ) comprises 25 items that fall under five subsections; emotional symptoms, conduct problems, hyperactivity/inattention, peer relationship problems, and prosocial behaviour. Each section is made up of 5 items rated on a three-point scale (‘Not true’, ‘Somewhat true’ or ‘Certainly true’). Analyses focused on items from the emotional subscale which include, “Often unhappy, down-hearted or tearful”; “Complains of headache/stomach ache”; “Many worries, often seems worried”; “Nervous or clingy in new situations, easily loses confidence”; and “Many fears, easily scared”. In line with the SDQ recommendations (www.sdqinfo.org), items were summed to generate an overall emotional score that ranges from 0 to 10, with mean imputation used for those with (≤2) of items missing. Further analyses focused on two core symptoms of depression and generalised anxiety, “Often unhappy, down-hearted or tearful” and “Many worries, often seems worried”.

The SDQ was administered and completed by mothers in ALSPAC at the approximate ages of 7, 10, 13, 16, and 25 years, as well as by self-report at age 25. This allowed validation against the depressive and anxiety diagnoses at similar timepoints.

#### 2.2.2 Diagnoses

Major Depressive Disorder (MDD) and Generalised Anxiety Disorder (GAD), as well as a range of other anxiety disorders (see Table 1 for details), were captured using the Development and Well-Being Assessment (DAWBA; Goodman et al., 2000). The DAWBA is a package of interviews, questionnaires, and rating techniques designed to generate psychiatric diagnoses. Versions of DAWBA related to the different disorders are available for parents or carers, teachers, and the individual. Parent-rated DAWBA questionnaire measures are available in ALSPAC at 7, 10, and 13 years, with self-report data available for depression at 15 and 25 years, and for GAD and other anxiety disorders at 15 years. Questions on MDD refer to the previous four weeks, while questions related to GAD refer to the last 6 months. Items specific to phobias refer to instances in the last month. It is worth noting that the gap between the DAWBA diagnoses and the SDQ assessments varied slightly across the different ages (see Supplementary).

**Table 1:** DAWBA diagnoses in ALSPAC

|  | Parent-reported |  |  | Self-reported |  |
| --- | --- | --- | --- | --- | --- |
|  | Age 7 | Age 10 | Age 13 | Age 15 | Age 25 |
| **Depressive disorder** |  |  |  |  |  |
| Major depressive disorder (MDD)** | ✓ | ✓ | ✓ | ✓ | ✓ |
| Depressive disorder NOS | ✓ |  |  |  |  |
| **Anxiety disorder** |  |  |  |  |  |
| Generalised Anxiety disorder (GAD)** | ✓ | ✓ | ✓ | ✓ |  |
| Separation anxiety disorder | ✓ | ✓ | ✓ |  |  |
| Specific phobia | ✓ | ✓ | ✓ | ✓ |  |
| Social phobia | ✓ | ✓ | ✓ | ✓ |  |
| Panic disorder |  |  |  | ✓ |  |
| Agoraphobia |  |  |  | ✓ |  |
| **Any anxiety disorder\*\*** | ✓ | ✓ | ✓ | ✓ |  |
| **Any emotional disorder** | ✓ | ✓ | ✓ | ✓ |  |
| **ADHD or any behavioural disorder \*\*** | ✓ | ✓ | ✓ | ✓ |  |

Note: \*\*Denotes the outcomes used in the current study. Any behavioural disorder includes Conduct Disorder (CD) and Oppositional Defiant Disorder (ODD). Depressive and anxiety diagnoses at ages 7, 10, and 13 were based on parent-reports only, and at 15 and 25 years were based on self-reports only. For ADHD or any behavioural disorder, these were all based on parent-reports.

The current study focused on whether individuals met the criteria for MDD, GAD, or any form of anxiety disorder assessed at each time point (see Table 1). In addition, as a secondary analysis to compare the performance of the emotional subscale for emotional and non-emotional disorders, we also used the parent DAWBA at 16 years to identify children without emotional disorders but who experienced other disorders including Attention Deficit Hyperactivity Disorder (ADHD) or any behavioural disorder (conduct disorder, oppositional defiant disorder). A full list of the DAWBA diagnoses included can be found in Table 1. It should be noted that there are minor differences in the anxiety disorders included at age 15 according to informant (see Table 1). Specifically, self-reports of ‘any anxiety disorder’ do not include separation anxiety disorder (which is included in the parent-rated ‘any anxiety disorder’ variable but they include the addition of panic disorder and agoraphobia). All specific diagnoses were based on algorithms generated according to DSM-IV, except at age 25 as this assessment was conducted after the release of the DSM-5 (APA, 2013). The computerised algorithm predicts the likelihood of a clinical rater assigning each child a diagnosis (see www.DAWBA.com for more information). The six-band computer prediction generated from these is then used to create a binary diagnosis variable according to whether the probability of a diagnosis is greater than 50%. This approach has been previously validated in a separate sample (Goodman et al., 2011). A senior clinical psychiatrist reviewed the diagnoses and DAWBA responses as part of the ALSPAC data collection process at age 7. As the DSM-5 no longer places Post-Traumatic Stress Disorder (PTSD) or Obsessive Compulsive Disorder (OCD) as anxiety disorders, these were removed in the generation of our ‘any anxiety’ disorder category at every age.

### 2.3 Analyses

All analyses were conducted in Stata version 17 and used Receiver Operating Characteristic (ROC) curve analyses. These enabled tests of the ability of the SDQ emotional subscale to distinguish between cases and non-cases of MDD, GAD, any form of anxiety disorder, and other disorder (ADHD or behavioural disorder not emotional disorder). A ROC curve was plotted (sensitivity vs 1-specificity), and the area under the curve (AUC) estimated. The area under the curve (AUC) was used to determine the diagnostic efficiency of the emotional subscale score in identifying individuals meeting the depressive or anxiety diagnostic criteria. An AUC value of 0.5 indicates performance at chance level, therefore values <0.7 are considered as having low test accuracy. Estimates between 0.7 and 0.9 are considered moderate, and those >0.9 as excellent when screening questionnaire performance against gold standard diagnostic measures (Henderson, 1993). It is worth noting that in some instances, such as prognostic prediction, different AUC values are sometimes considered relevant.

Sensitivity (the ability of the screening questionnaire to correctly identify those with a diagnosis) and specificity (the ability of the screening questionnaire to correctly identify those without a diagnosis) estimates were derived from the ROC curve analyses and used to explore optimum cut-off scores for the full emotional subscale (range 0 to 10). Optimum cut-points for diagnosis were selected as those that best balanced sensitivity and specificity according to maximal Youden Index (sensitivity + specificity – 1) (Fluss et al, 2005). These tests of sensitivity and specificity form part of the Standards for Reporting Diagnostic Accuracy (STARD) initiative (Bossuyt et al., 2003; Bossuyt et al., 2015).

Positive predictive values (PPV) and negative predictive values (NPV) were calculated for the optimal cut-point to show the probability that those exceeding the cut-point have a diagnosis, and the probability that those not meeting the cut-point do not have a diagnosis, respectively. These predictive values are dependent on sample prevalence rates. Following this, analyses investigated two individual items from the emotional subscale to determine whether a) the low mood item was better at discriminating MDD compared to the worry item, and b) whether the worry item was better at discriminating GAD compared to the low mood item. This was done using Stata’s roccomp function (Cleves, 2002).

#### 2.3.1 Sensitivity analyses

Sensitivity analyses explored the ability of the emotional subscale score, and two individual items in detecting cases of MDD, GAD, and any anxiety disorder among males and females separately. These analyses were also run using Stata’s roccomp function which allows comparison of AUC values by sex.

#### 2.3.2 Missing data

To examine whether any differences in findings across time points could be reflective of different patterns of missing data across ages rather than developmental differences, supplementary analyses used multiple imputation. Multiple imputation by chained equations (MICE) was used to impute missing diagnostic and SDQ data at each time point for individuals who had data at least once on the SDQ, and who completed at least one DAWBA assessment (n=9,241). One hundred imputed datasets were produced, and estimates were pooled across these 100 datasets. All analysis variables were included in the imputation models alongside auxiliary variables previously shown to predict missingness in the ALSPAC cohort (see Supplementary Table 3 for full list). Imputation was conducted using Predictive Mean Matching (PMM) (Royston, 2005). This matches the predictive mean distance of incomplete observations with the complete observations. Datasets were imputed using the ‘mi estimate’ and ‘eroctab’ commands in Stata which fit a model to each of the imputed data sets and pool individual results using Rubin’s combination rules (White et al., 2011).

## 3. Results

### 3.1 Descriptives

The internal consistency of SDQ-E scores averaged 0.69, which is slightly above previous findings of 0.67 (Goodman 2001), and there was some evidence of improvement with age (α=0.63 at 7 years, α=0.68 at 10 years, α=0.67 at 13 years, α=0.71 at 16 years, α=0.77 at 25 years). Total scores on the emotional subscale remained relatively stable across childhood and adolescence, but were somewhat higher in young adulthood (particularly for self-reports) (Supplementary table 1). Females scored more highly than males at all time points (see Supplementary Table 2), as anticipated based on prevalence rates of emotional disorders between sexes (Thapar et al., 2022).

For the diagnoses in our study, the prevalence of both depressive and anxiety disorders varied across development, with both MDD and GAD most common at the oldest age assessed (25 years for MDD and 15 years for GAD), and least common at 7 years (see Supplementary Table 1). Varying rates of diagnosis were noted across males and females according to age and disorder (see Table 2). Males typically had higher rates of depression and anxiety than females in childhood, but the prevalence was greater for females compared to males in adolescence.

**Table 2:**
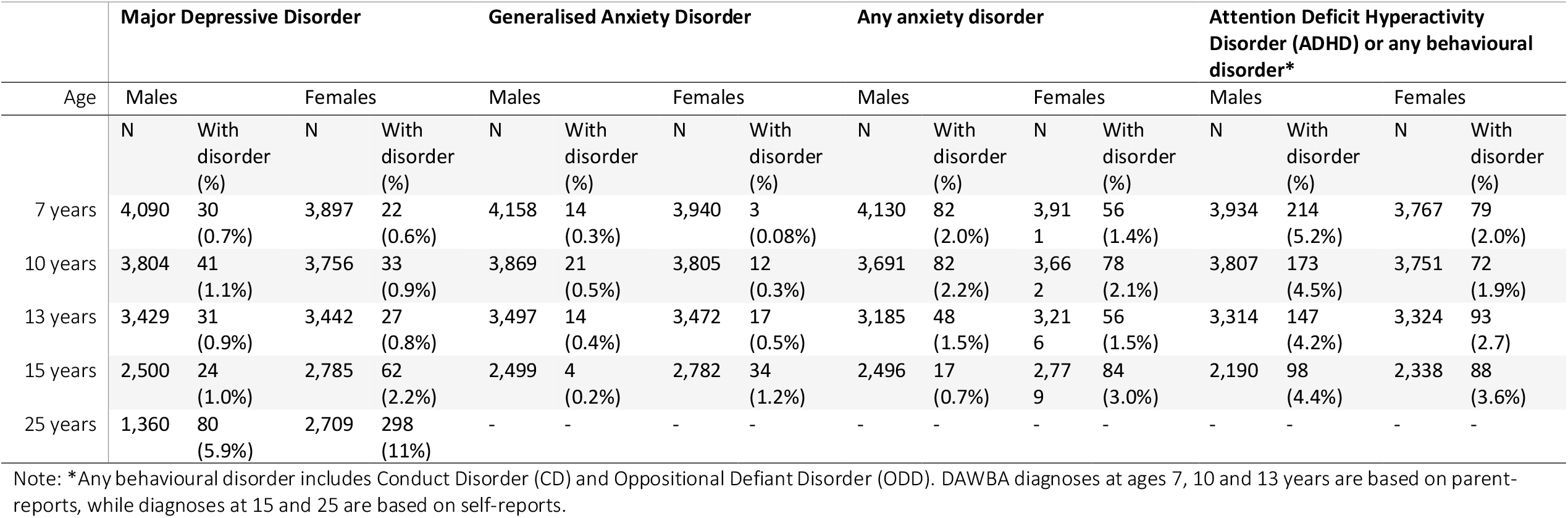
Descriptives of DAWBA diagnoses by sex

### 3.2 Validation of the emotional subscale

The Receiver Operating Characteristic (ROC) curves are shown in Figure 1 for MDD and GAD (see Supplementary Figures 1 and 2 for any anxiety and ADHD/any behavioural disorder outcomes). These analyses showed that the emotional subscale had moderate accuracy in discriminating between cases and non-cases of MDD (AUC range = 0.67-0.85), and high accuracy in discriminating cases of GAD (AUC range = 0.80-0.93) and any anxiety disorder (0.74-0.83) from non-cases (see Table 3). Accuracy tended to be higher for GAD and any anxiety disorder across all time points compared to MDD. No clear pattern was observed across development for any of the disorder outcomes and there were overlapping confidence intervals across all time points for each disorder, suggesting no differences by age. In particular, comparing parent reports (available across age) there was no evidence for any consistent increase or decrease in accuracy with age. As expected, and providing evidence of discriminant validity, the SDQ-E performed less well at identifying other disorders (ADHD or behavioural) in the absence of an emotional disorder (AUC range = 0.61-0.70). Results from the imputed dataset revealed largely consistent findings (Supplementary Table 4).

**Table 3:** Discrimination of those with versus without DAWBA diagnoses for the SDQ emotional subscale and depressive and worry items

| Age | Emotional subscale<br>AUC (95% CI) | Depressive item<br>AUC (95% CI) | Worry item<br>AUC (95% CI) | Emotional subscale versus depressive item<br>$\chi^2_{(1)}$, p-value | Emotional subscale versus worry item<br>$\chi^2_{(1)}$, p-value | Depressive item versus worry item<br>$\chi^2_{(1)}$, p-value |
| --- | --- | --- | --- | --- | --- | --- |
| **Major Depressive Disorder** |  |  |  |  |  |  |
| 7 years | 0.76 (0.69, 0.84) | 0.71 (0.63, 0.78) | 0.73 (0.65, 0.81) | 1.93, 0.17 | 1.17, 0.28 | 0.23, 0.63 |
| 10 years | 0.76 (0.71, 0.82) | 0.66 (0.60, 0.73) | 0.75 (0.69, 0.81) | 15.04, **<0.001** | 1.05, 0.30 | 6.86, **<0.01** |
| 13 years | 0.85 (0.79, 0.90) | 0.76 (0.69, 0.83) | 0.73 (0.66, 0.81) | 8.74, **<0.01** | 16.66, **<0.001** | 0.48, 0.49 |
| 15/16 years | 0.67 (0.60, 0.74) | 0.61 (0.54, 0.67) | 0.66 (0.59, 0.73) | 3.92, **<0.01** | 0.21, 0.64 | 1.68, 0.19 |
| 25 years | 0.72 (0.68, 0.76) | 0.68 (0.65, 0.72) | 0.67 (0.63, 0.70) | 8.02, **<0.01** | 22.33, **<0.001** | 1.45, 0.23 |
| 25 years (self) | 0.85 (0.83, 0.87) | 0.86 (0.84, 0.88) | 0.74 (0.72, 0.76) | 1.72, 0.19 | 166.4, **<0.001** | 105.0, **<0.001** |
| **Generalised Anxiety Disorder** |  |  |  |  |  |  |
| 7 years | 0.91 (0.82, 0.99) | 0.79 (0.65, 0.93) | 0.83 (0.72, 0.95) | 4.41, **<0.05** | 3.84, **<0.05** | 0.27, 0.60 |
| 10 years | 0.87 (0.79, 0.94) | 0.75 (0.65, 0.85) | 0.82 (0.73, 0.90) | 8.84, **<0.01** | 3.24, 0.07 | 2.16, 0.14 |
| 13 years | 0.93 (0.90, 0.97) | 0.81 (0.72, 0.90) | 0.87 (0.80, 0.93) | 10.35, **<0.001** | 7.16, **<0.01** | 1.52, 0.22 |
| 15/16 years | 0.80 (0.74, 0.87) | 0.75 (0.64, 0.85) | 0.72 (0.61, 0.82) | 2.69, 0.10 | 5.10, **<0.05** | 0.42, 0.52 |
| **Any anxiety disorder** |  |  |  |  |  |  |
| 7 years | 0.81 (0.77, 0.85) | 0.63 (0.58, 0.68) | 0.71 (0.66, 0.76) | 66.64, **<0.001** | 31.20, **<0.001** | 8.08, **<0.01** |
| 10 years | 0.79 (0.74, 0.83) | 0.67 (0.63, 0.71) | 0.70 (0.66, 0.75) | 39.68, **<0.001** | 28.82, **<0.001** | 2.00, 0.16 |
| 13 years | 0.83 (0.78, 0.87) | 0.69 (0.64, 0.75) | 0.75 (0.69, 0.80) | 36.34, **<0.001** | 14.76, **<0.001** | 3.68, **<0.05** |
| 15/16 years | 0.74 (0.68, 0.80) | 0.65 (0.59, 0.71) | 0.63 (0.56, 0.70) | 13.43, **<0.001** | 18.78, **<0.001** | 0.42, 0.52 |
| **Attention Deficit Hyperactivity Disorder or any behavioural disorder\*** |  |  |  |  |  |  |
| 7 years | 0.61 (0.58, 0.65) | 0.61 (0.58, 0.64) | 0.57 (0.54, 0.60) | 0.10, 0.75 | 6.68, **<0.01** | 4.94, **<0.05** |
| 10 years | 0.66 (0.62, 0.69) | 0.63 (0.60, 0.66) | 0.61 (0.58, 0.65) | 2.31, 0.13 | 6.51, **<0.01** | 0.80, 0.37 |
| 13 years | 0.64 (0.60, 0.68) | 0.61 (0.57, 0.64) | 0.61 (0.57, 0.65) | 5.91, **<0.05** | 4.51, **<0.05** | 0.11, 0.74 |
| 15/16 years | 0.70 (0.65, 0.75) | 0.65 (0.60, 0.70) | 0.63 (0.58, 0.68) | 8.45, **<0.01** | 11.32, **<0.001** | 0.10, 0.76 |

Note: Any behavioural disorder includes Conduct Disorder (CD) and Oppositional Defiant Disorder (ODD). SDQ assessments are based on the concurrent age of the diagnosis, however there is a some gap between assessments (see supplementary). All SDQ assessments are based on parent-reports unless stated otherwise. Diagnoses at ages 7, 10 and 13 years are based on parent-reports, while diagnoses at 15 and 25 years are based on self-reports.

**Figure 1:**
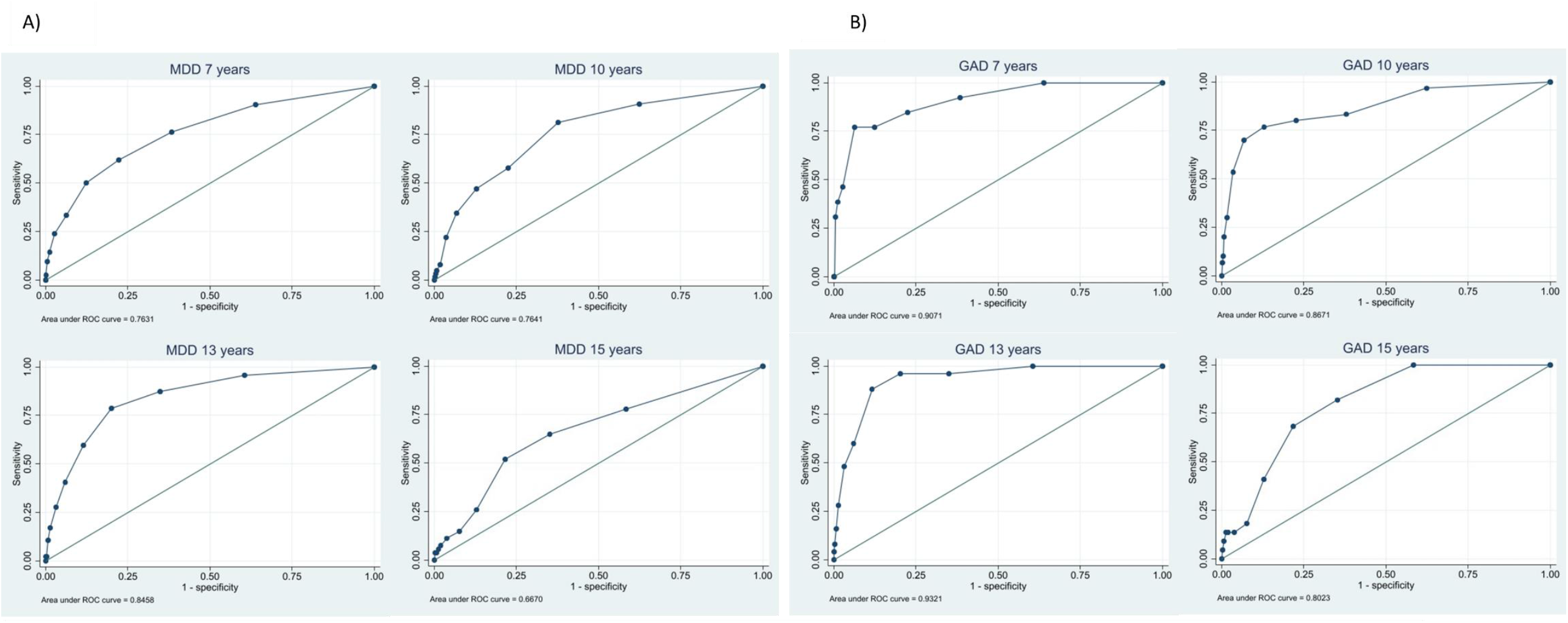
ROC analyses for emotional subscale predicting (A) Major Depressive Disorder and (B) Generalised Anxiety Disorder across development.

Sensitivity and specificity values for all possible SDQ emotional subscale cut-points are reported in Supplementary Tables 5 to 7. Optimal cut points, which balanced sensitivity and specificity, were generally lower for MDD diagnoses in childhood and adolescence compared to those for GAD. For MDD, a score of 3 or higher captured between 52% and 81% of MDD diagnoses across development, while a score of 4 or 5 for GAD captured between 77% and 88% of cases across development (see Supplementary Table 8).

When investigating the two individual items (low mood and general worry), these did not perform as well as the full emotional subscale in detecting depressive or anxiety diagnoses respectively (Table 3). The low mood item demonstrated greater accuracy than the general worry item in detecting MDD at 25 years when self-reported. The low mood and worry items showed similar accuracy in detecting cases of GAD, but the worry item performed better for detecting cases of any anxiety disorder at 7 and 13 years (see Table 3).

### 3.3 Sensitivity analyses

We did not find consistent sex differences in the accuracy of the SDQ-E in identifying cases of MDD, GAD or other anxiety disorders (see Table 4 and also Supplementary Tables 9 and 10).

**Table 4:** Discrimination of those with versus without DAWBA diagnoses for the emotional subscale by sex

| Age | Major Depressive Disorder |  |  | Generalised Anxiety Disorder |  |  | Any anxiety disorder |  |  | Attention Deficit Hyperactivity Disorder (ADHD) or any behavioural disorder* |  |  |
| --- | --- | --- | --- | --- | --- | --- | --- | --- | --- | --- | --- | --- |
| | Males<br>AUC<br>(95% CI) | Females<br>AUC<br>(95% CI) | Diff<br>$\chi^2_{(1)}$,<br>p-value | Males<br>AUC<br>(95% CI) | Females<br>AUC<br>(95% CI) | Diff<br>$\chi^2_{(1)}$, p-value | Males<br>AUC<br>(95% CI) | Females<br>AUC<br>(95% CI) | Diff<br>$\chi^2_{(1)}$, p-value | Males<br>AUC<br>(95% CI) | Females<br>AUC<br>(95% CI) | Diff<br>$\chi^2_{(1)}$, p-value |
| 7 years | 0.80<br>(0.70, 0.89) | 0.71<br>(0.57, 0.85) | 1.08,<br>0.30 | 0.89<br>(0.79, 0.99) | 0.98<br>(0.95, 1.00) | 2.68,<br>0.10 | 0.81<br>(0.76, 0.87) | 0.81<br>(0.75, 0.88) | 0.00,<br>0.99 | 0.61<br>(0.57, 0.65) | 0.63<br>(0.56, 0.70) | 0.20,<br>0.65 |
| 10 years | 0.77<br>(0.69, 0.85) | 0.76<br>(0.67, 0.84) | 0.04,<br>0.84 | 0.89<br>(0.80, 0.98) | 0.83<br>(0.69, 0.97) | 0.41,<br>0.52 | 0.82<br>(0.76, 0.88) | 0.75<br>(0.69, 0.81) | 2.64,<br>0.10 | 0.67<br>(0.63, 0.71) | 0.64<br>(0.57, 0.72) | 0.42,<br>0.52 |
| 13 years | 0.86<br>(0.77, 0.94) | 0.83<br>(0.76, 0.90) | 0.20,<br>0.66 | 0.94<br>(0.88, 0.99) | 0.93<br>(0.90, 0.96) | 0.03,<br>0.86 | 0.86<br>(0.80, 0.91) | 0.80<br>(0.73, 0.87) | 1.55,<br>0.21 | 0.64<br>(0.59, 0.70) | 0.67<br>(0.61, 0.73) | 0.46,<br>0.50 |
| 15/16 years | 0.64<br>(0.49, 0.79) | 0.65<br>(0.56, 0.74) | 0.02,<br>0.89 | 0.89<br>(0.88, 0.91) | 0.74<br>(0.66, 0.83) | 11.63,<br>**<0.001** | 0.54<br>(0.37, 0.70) | 0.74<br>(0.68, 0.80) | 5.09,<br>**<0.05** | 0.66<br>(0.60, 0.73) | 0.78<br>(0.72, 0.84) | 6.00,<br>**<0.01** |
| 25 years | 0.68<br>(0.59, 0.76) | 0.72<br>(0.68, 0.77) | 1.05,<br>0.31 | - | - | - | - | - | - | - | - | - |
| 25 years (self) | 0.88<br>(0.85, 0.91) | 0.83<br>(0.81, 0.85) | 5.32,<br>**<0.05** | - | - | - | - | - | - | - | - | - |

Note: \*Any behavioural disorder includes Conduct Disorder (CD) and Oppositional Defiant Disorder (ODD). All DAWBA diagnoses at ages 7, 10 and 13 years are based on parent-reports, while diagnoses at 15 and 25 are based on self-reports. All SDQ assessments based on self-reports unless stated otherwise.

## 4. Discussion

This study aimed to examine the validity of the emotional subscale of the SDQ in identifying depression and anxiety diagnoses across development. Our findings provide validation that the subscale can be used to distinguish between those meeting the diagnostic criteria for MDD, GAD, or any other anxiety disorder from those who do not. This was the case for disorders across all ages, and for both males and females. Such findings provide important validation for a time-efficient tool that can be used by researchers studying depressive or anxiety disorders across development.

The accuracy of the emotional subscale for distinguishing between cases and non-cases was high for GAD and anxiety disorders, and moderate for MDD, in line with previous research on children (Silva et al., 2015). Our findings extend these to provide the first evidence that validity is largely stable across time for both depression and anxiety, with AUC values ranging from 0.72 to 0.85 for MDD, from 0.80 to 0.93 across development for GAD, and from 0.74 to 0.83 for any anxiety disorder. Accuracy was also stable for both sexes, suggesting that while the presentation of symptoms may vary with age and sex, the items within the emotional subscale capture a similar construct that can be examined fairly across development and for males and females.

The AUC values in the present study balanced specificity and sensitivity. This is the typical approach used in research whereby an individual is categorised as having a disorder in one step. False positives and false negatives in this instance are therefore equally undesirable. In a clinical setting, a two-step process is often used such that a questionnaire or interview is followed by further assessments. These initial assessments may be more accepting of false positives and favour a cut-point that prioritises sensitivity over specificity. During the second diagnostic step, clinicians may then instead favour specificity over sensitivity (Silva et al., 2015). Variable cut-points may therefore be required depending on the rationale for using the instrument, as outlined in the STARD statement (Bossuyt et al., 2015). Our findings provide insight into specific cut-points recommended for the SDQ-E to detect depression and anxiety disorders across development.

Current recommendations for the SDQ-E suggest that in childhood and adolescence, parent-rated emotional scores of 5 or 6 capture the top 10% of ‘abnormal’ scores in the population, while a score of 4 captures those with slightly raised or ‘borderline’ symptoms. Our results suggest a lower cut-point may be needed to capture clinical symptoms of MDD in childhood and adolescence when using parent-ratings. In particular, findings revealed that a score of 3 or higher captured between 52% and 81% of MDD diagnoses across development, compared to a score of 4 or 5 for GAD, which captured between 77% and 88% of cases across development. When using self-reports to capture MDD at 25 years, a score of 5 was the optimal cut-off point (see Supplementary Table 10 for more detail). Note that the optimal cut-point will also depend on the setting and whether it is more desirable to avoid false positives or false negatives.

### 4.1 Strengths and limitations

By using a longitudinal design, our study overcomes previous limitations related to the generalisability of results from childhood to adolescence and beyond. This is crucial for determining whether the same construct can be used in research focused on depressive and anxiety disorders across development. Several limitations of this study, however, should be noted.

First, there was a gap of 9-13 months between the SDQ and DAWBA assessments at each age. Estimates of the validity of the SDQ-E are therefore likely conservative. This may present a particular issue for MDD which is often episodic, and may explain why the SDQ-E appeared to perform more poorly for MDD compared to anxiety. Indeed, at 25 years where the age gap between the completion of the SDQ and DAWBA was minimal, the accuracy for depression was higher. It is also possible that accuracy was lower during late adolescence because of differences in informant. At 15/16 years, parent and self-report were available for the SDQ-E and DAWBA diagnoses respectively. All analyses prior to this relied on parent-reports for both the SDQ-E and DAWBA assessment. Future validation studies should attempt to replicate our findings using both parent and adolescent self-reports of diagnoses and the SDQ-E. This multi-informant approach is commonly used in practice, and it would help to ensure findings do not underreport the frequency or severity of emotional problems, and would minimise bias that could arise from using different informants for different assessments.

Second, future research should also seek to use clinical diagnoses. As per most epidemiological studies on depression and anxiety, diagnoses in the current study were not generated using clinical interviews with trained interviewers, meaning information available to derive diagnoses was more limited.

Other considerations relate to the prevalence and comorbidity of emotional disorders. The overall prevalence of MDD, GAD, and any anxiety diagnosis was relatively low, likely due to the young age and stringent DAWBA assessments. Co-morbidity of depressive and anxiety disorders were also common, and there were varying rates noted for females and males (Supplementary Table 11). This aligns with previous research which has shown that compared to adolescents who are not depressed, those with a depression diagnosis are six to 12 times more likely to also have anxiety (Costello et al., 2006). Rates of comorbidity may have altered the accuracy of the subscale in detecting cases of anxiety or depression, particularly as more individuals had GAD with MDD compared to having MDD with anxiety. Further sensitivity analyses removing comorbid cases would have been underpowered, but should be considered in larger validation studies.

Finally, ALSPAC like other prospective birth cohorts experienced non-trivial participant drop-out over time. Previous work has shown that children with increased risk for mental health problems and more disadvantaged families were less likely to participate in the study in childhood and adolescence (Boyd et al, 2013; Martin et al, 2016). However, sensitivity analyses accounting for attrition across all follow-up time points through multiple imputation showed closely similar findings.

### 4.2 Conclusion

Overall, our findings suggest the emotional subscale of the SDQ is an appropriate measure to determine the risk of emotional disorders like depression and anxiety among males and females across childhood and adolescence, and in adulthood for depression. This provides important validation for the use of this subscale in monitoring longitudinal changes in emotional problems across development within research. Retaining the same measure to study the developmental course of depressive and anxiety disorders is especially important when studying changes in incidence and outcomes as it helps to ensure differences are less likely a result of measurement changes. Our findings could therefore aid the assessment and monitoring of those at risk of depressive or anxiety disorders to both reduce their prevalence and associated long-term effects.

## Data Availability

The University of Bristol owns the ALSPAC resource. Data can be made available on request to the ALSPAC Executive. Full instructions for applying for data access can be found here: http://www.bristol.ac.uk/alspac/researchers/access/.The ALSPAC study website contains details of all the data that are available (http://www.bristol.ac.uk/alspac/researchers/our-data/).

http://www.bristol.ac.uk/alspac/researchers/access/

## Acknowledgements

We are extremely grateful to all the families who took part in this study, the midwives for their help in recruiting them, and the whole ALSPAC team, which includes interviewers, computer and laboratory technicians, clerical workers, research scientists, volunteers, managers, receptionists and nurses.

This work was supported by the Wolfson Centre for Young People’s Mental Health. The UK Medical Research Council and Wellcome (Grant ref: 102215/2/13/2) and the University of Bristol provide core support for ALSPAC. A comprehensive list of grants funding is available on the ALSPAC website (http://www.bristol.ac.uk/alspac/external/documents/grant-acknowledgements.pdf). This publication is the work of the authors and Jessica Armitage and Stephan Collishaw will serve as guarantors for the contents of this paper.

## Author contributions

JMA, LR, and SC conceptualised the study and JMA wrote the original draft. JMA performed all statistical analyses, and LR and SC provided supervision. All authors contributed to the interpretation of data and reviewed/edited the manuscript.

