## Supplementary material for "Validation of the Strengths and Difficulties Questionnaire (SDQ) emotional subscale in assessing depression and anxiety across development"

- Supplementary Table 1: Descriptives of samples included
- Supplementary Table 2: Descriptives of Strengths and Difficulties Questionnaire by sex
- Supplementary Table 3: Variables included in multiple imputation
- Supplementary Table 4: Discrimination of those with versus without DAWBA diagnoses for the emotional subscale and depressive and worry items using imputed dataset (n=9,241)
- Supplementary Table 5: Sensitivity and specificity of the emotional subscale cutoff-points across development compared against Major Depressive Disorder Diagnoses
- Supplementary Table 6: Sensitivity and specificity of the emotional subscale cutoff-points across development compared against Generalised Anxiety Disorder Diagnoses
- Supplementary Table 7: Sensitivity and specificity of the emotional subscale cutoff-points across development compared against any anxiety disorder diagnoses
- Supplementary Table 8: Accuracy of identifying those meeting diagnostic criteria by optimal SDQ subscale cut-point
- Supplementary Table 9: Discrimination of those with versus without DAWBA diagnoses for the depressive item by sex
- Supplementary Table 10: Discrimination of those with versus without DAWBA diagnoses for the worry item by sex
- Supplementary Table 11: Comorbidity of DAWBA Depressive and Anxiety disorders by sex

| **Supplementary Table 1: Descriptives of samples included** | | | | | | | | | | | | | | | | |
| --- | --- | --- | --- | --- | --- | --- | --- | --- | --- | --- | --- | --- | --- | --- | --- | --- |
| **Strengths and Difficulties Questionnaire (SDQ)** | | | | | | | | **Development and Well-Being Assessment (DAWBA) Diagnoses** | | | | | | | | |
| **Age** | **Emotional subscale**  **(range 0-10)** | | | **Depressive item**  **(range 0-2)** | | **Worry item**  **(range 0-2)** | | **Age** | **Major Depressive disorder** | | **Generalised Anxiety Disorder** | | **Any anxiety disorder** | | **Attention Deficit Hyperactivity Disorder (ADHD) or any behavioural disorder*** | |
|  | N | Mean (SD) | α | N | Mean (SD) | N | Mean (SD) |  | N | With disorder (%) | N | With disorder (%) | N | With disorder (%) | N | With disorder (%) |
| 7  years | 8,312 | 1.51 (1.67) | 0.63 | 8,298 | 0.17 (0.41) | 8,256 | 0.29 (0.52) | 7 years | 7987 | 52 (0.7%) | 8,098 | 17 (0.2%) | 8,041 | 138 (1.7%) | 8,102 | 293 (3.6%) |
| 10 years | 7,956 | 1.52 (1.77) | 0.68 | 7,905 | 0.18 (0.42) | 7,833 | 0.30 (0.52) | 10 years | 7,560 | 74 (1.0%) | 7,674 | 33 (0.4%) | 8,063 | 160 (2.2%) | 7,558 | 245 (3.3%) |
| 13 years | 6,970 | 1.44 (1.72) | 0.67 | 6,936 | 0.18 (0.43) | 6,914 | 0.29 (0.52) | 13 years | 6,871 | 58 (0.8%) | 6,969 | 31 (0.4%) | 6,401 | 104 (1.6%) | 6,638 | 240 (3.5%) |
| 16 years | 5,590 | 1.50 (1.86) | 0.71 | 5,558 | 0.19 (0.46) | 5,504 | 0.40 (0.60) | 15 years  (self) | 5,293 | 86 (1.6%) | 5,289 | 38 (0.7%) | 5,275 | 101 (1.9%) | 4,533 | 186 (4.0%) |
| 25 years | 4,393 | 1.90 (2.23) | 0.77 | 4,387 | 0.26 (0.55) | 4,319 | 0.54 (0.68) | 25 years  (self) | 4,074 | 378 (9.3%) | - | - | - | - | - | - |
| 25 years  (self) | 4,309 | 3.44 (2.49) | 0.75 | 4,305 | 0.49 (0.66) | 4,295 | 1.10 (0.77) | - | - | - | - | - | - | - | - | - |
| Note: α = Cronbach’s alpha. *Any behavioural disorder includes Conduct Disorder (CD) and Oppositional Defiant Disorder (ODD).  Sample sizes for the individual items are lower than the total subscale as this used a mean imputation procedure for those missing ≤2 items. All SDQ assessments are based on parent-reports unless stated otherwise. DAWBA diagnoses at ages 7, 10 and 13 years are based on parent-reports, while diagnoses at 15 and 25 are based on self-reports. At 7 years, the mean age during assessment of the SDQ was 81 months, and 91 months for the DAWBA diagnoses. At age 10, the mean age of assessment for the SDQ was 115 months, and 128 months for the DAWBA. At 13 years mean ages were 157 months and 166 months respectively, and at 15/16 years, the SDQ was parent-rated at 198 months, while the DAWBA diagnoses were self-reported at 185 months. | | | | | | | | | | | | | | | | |

| **Supplementary Table 2: Descriptives of Strengths and Difficulties Questionnaire by sex** | | | | | | | | | | | | | | | |
| --- | --- | --- | --- | --- | --- | --- | --- | --- | --- | --- | --- | --- | --- | --- | --- |
|  | **Emotional subscale (range 0-10)** | | | | | **Depressive item (range 0-2)** | | | | | **Worry item (range 0-2)** | | | | |
| **Age** | **Males** | | **Females** | | **Diff *** | **Males** | | **Females** | | **Diff *** | **Males** | | **Females** | | **Diff *** |
|  | N | Mean (SD) | N | Mean  (SD) | Estimate  (95% CI) | N | Mean (SD) | N | Mean  (SD) | Estimate  (95% CI) | N | Mean (SD) | N | Mean  (SD) | Estimate  (95% CI) |
| 7 years | 4,265 | 1.43 (1.66) | 4,047 | 1.58 (1.68) | **0.15**  (0.08, 0.22) | 4,258 | 0.16 (0.41) | 4,040 | 0.18 (0.41) | 0.02  (-0.004, 0.03) | 4,231 | 0.29 (0.53) | 4,025 | 0.29 (0.51) | 0.00  (-0.03, 0.02) |
| 10 years | 4,015 | 1.39 (1.71) | 3,941 | 1.66 (1.82) | **0.27**  (0.20, 0.35) | 3,986 | 0.16 (0.40) | 3,919 | 0.20 (0.44) | **0.04**  (0.02, 0.05) | 3,957 | 0.29 (0.52) | 3,876 | 0.30 (0.52) | 0.01  (-0.01, 0.04) |
| 13 years | 3,471 | 1.24 (1.61) | 3,499 | 1.63 (1.79) | **0.39**  (0.31, 0.47) | 3,465 | 0.15 (0.40) | 3,480 | 0.20 (0.46) | **0.05**  (0.03, 0.07) | 3,443 | 0.26 (0.50) | 3,471 | 0.31 (0.53) | **0.04**  (0.02, 0.07) |
| 16 years | 2,676 | 1.08 (1.55) | 2,878 | 1.87 (2.03) | **0.79**  (0.69, 0.88) | 2,676 | 0.11 (0.37) | 2,882 | 0.26 (0.52) | **0.15**  (0.12, 0.17) | 2,641 | 0.32 (0.56) | 2,863 | 0.46 (0.63) | **0.14**  (0.11, 0.17) |
| 25 years | 2,042 | 1.50 (1.94) | 2,351 | 2.25 (2.40) | **0.74**  (0.61, 0.87) | 2,041 | 0.20 (0.49) | 2,346 | 0.32 (0.59) | **0.12**  (0.08, 0.15) | 2,001 | 0.46 (0.64) | 2,318 | 0.60 (0.71) | **0.14**  (0.10, 0.18) |
| 25 years  (self) | 1,455 | 2.62 (2.21) | 2,849 | 3.87 (2.52) | **1.25**  (1.10, 1.40) | 1,454 | 0.40 (0.64) | 2,846 | 0.53 (0.67) | **0.13**  (0.09, 0.17) | 1,451 | 0.85 (0.76) | 2,839 | 1.23 (0.74) | **0.38**  (0.33, 0.43) |
| Note: *Difference represents the degree to which females score more highly than males at that age point. Those in bold are statistically different.  Sample sizes for the individual items are lower than the total subscale as this used a mean imputation procedure for those missing <2 items. All SDQ assessments are based on parent-reports unless stated otherwise. At 7 years, the mean age during assessment of the SDQ was 81 months, and 91 months for the DAWBA diagnoses. At age 10, the mean age of assessment for the SDQ was 115 months, and 128 months for the DAWBA. At 13 years the SDQ and DAWBA were assessed at mean ages 157 months and 166 months respectively, and at 15/16 years, the SDQ was assessed at 198 months, while the DAWBA diagnoses were self-reported at 185 months. | | | | | | | | | | | | | | | |

| **Supplementary Table 3: Variables included in multiple imputation** | | | | |
| --- | --- | --- | --- | --- |
| **Variable** | **Age at assesment** | **Respondent** | **Item** | **Item response** |
| SDQ emotional problems subscale items | 7, 10, 13, 16, and 25 years | Mother | Strengths and Difficulties Questionnaire (SDQ) | ‘Not true’/‘Somewhat true’/‘Certainly true’ |
| Participant MDD, GAD, any anxiety, and any ADHD or behavioural diagnosis | 7, 10, 13, 15, and 25 years  25 years | Mother  Self | Development and Well-Being Assessment (DAWBA) | With disorder/Without disorder |
| Family ethnicity | 32 weeks gestation | Mother | “How would you describe the race or ethnic group of yourself/your partner?” | 8 response options. |
| Mother’s age at first pregnancy | 18 weeks gestation | Mother | “How old were you when you became pregnant for the very first time?” | Age in years. |
| Mother marital status | 32 weeks gestation | Mother | “What is your present marital status? | 6 response options including “Married” and “Separated” |
| Mother educational qualifications | 32 weeks gestation | Mother | “What educational qualifications do you/your partner have?” | List of qualifications, respondent required to tick all that apply |
| Mother home ownership status | 8 months | Mother | “Do you currently live in..” | 6 response options including “Mortgaged” and “Rented from private landlord” |
| Mother economic status | 32 weeks gestation | Mother | “What is the present employment situation of yourself?” | 11 response options including “Working for an employer full time”, “Self-employed”, “In full time education” and “Looking after home/family” |
| Mother smoking during pregnancy | 8 weeks | Mother | Did you smoke regularly in the last 2 months of pregnancy and since having the baby? | Options include no/ yes (cigarettes/pipe/cigar/other) |
| Mother mental health during pregnancy | 8 months | Mother | Edinburgh Post-natal Depression Score | 10 item questionnaire rated. Those scoring 13 or above are considered to be suffering from depressive disorder |

| **Supplementary Table 4: Discrimination of those with versus without DAWBA diagnoses for the emotional subscale and depressive and worry items using imputed dataset (n=9,241)** | | | | |
| --- | --- | --- | --- | --- |
| **Age (years)** | **Emotional subscale**  **AUC (95% CI)** | | | |
|  | **Major Depressive Disorder** | **Generalised Anxiety Disorder** | **Any anxiety disorder** | **Attention Deficit Hyperactivity Disorder (ADHD) or any behavioural disorder*** |
| 7 years | 0.77 (0.70, 0.84) | 0.89 (0.78, 0.99) | 0.81 (0.77, 0.85) | 0.61 (0.58, 0.65) |
| 10 years | 0.76 (0.70, 0.82) | 0.86 (0.78, 0.94) | 0.78 (0.74, 0.82) | 0.65 (0.61, 0.69) |
| 13 years | 0.82 (0.76, 0.88) | 0.91 (0.85, 0.97) | 0.82 (0.77, 0.87) | 0.64 (0.60, 0.68) |
| 15/16 years | 0.68 (0.61, 0.74) | 0.74 (0.65, 0.84) | 0.71 (0.66, 0.77) | 0.70 (0.66, 0.74) |
| 25 years | 0.74 (0.70, 0.77) | - | - | **-** |
| 25 years (self) | 0.84 (0.82, 0.86) | - | - | - |
| Note: Any behavioural disorder includes Conduct Disorder (CD) and Oppositional Defiant Disorder (ODD). SDQ assessments are based on the concurrent age of the diagnosis, however there is a gap between assessments. All SDQ assessments are based on parent-reports unless stated otherwise. Diagnoses at ages 7, 10 and 13 years are based on parent-reports, while diagnoses at 15 and 25 years are based on self-reports. | | | | |

| **Supplementary Table 5: Sensitivity and specificity of the emotional subscale cutoff-points across development compared against Major Depressive Disorder Diagnoses** | | | | | | | | | | | | |
| --- | --- | --- | --- | --- | --- | --- | --- | --- | --- | --- | --- | --- |
| Cut-point | Major Depressive Disorder at 7 years | | Major Depressive Disorder at 10 years | | Major Depressive Disorder at 13 years | | Major Depressive Disorder at 15/16 years | | Major Depressive Disorder at 25 years (based on parent-rated SDQ) | | Major Depressive Disorder at 25 years (based on self-rated SDQ) | |
|  | Sensitivity | Specificity | Sensitivity | Specificity | Sensitivity | Specificity | Sensitivity | Specificity | Sensitivity | Specificity | Sensitivity | Specificity |
| ≥ 1 | 90.48% | 36.14% | 90.63% | 37.56% | 95.74% | 39.50% | 79.25% | 41.72% | 86.60% | 39.15% | 100.00% | 12.88% |
| ≥ 2 | 76.19% | 61.69% | **81.25%**  **PPV=2%** | **62.33%**  **NPV=>99%** | 87.23% | 65.13% | 66.04% | 65.14% | 73.71% | 56.98% | 98.94% | 28.77% |
| ≥ 3 | **61.90%**  **PPV=2%** | **77.73%**  **NPV=>99%** | 57.81% | 77.48% | **78.72%**  **PPV=3%** | **79.95%**  **NPV=>99%** | **52.83%**  **PPV=3%** | **78.57%**  **NPV=>99%** | **59.28%**  **PPV=15%** | **73.06%**  **NPV=96%** | 94.97% | 45.74% |
| ≥ 4 | 50.00% | 87.72% | 46.88% | 87.14% | 59.57% | 88.51% | 26.42% | 87.17% | 47.42% | 83.24% | 90.21% | 60.46% |
| ≥ 5 | 33.33%  PPV=3% | 93.79%  NPV=99% | 34.38%  PPV=5% | 93.20%  NPV=99% | 40.43%  PPV=5% | 94.07%  NPV=99% | 15.09%  PPV=3% | 92.49%  NPV=99% | 38.14%  PPV=24% | 89.90%  NPV=95% | **83.07%**  **PPV=24%** | **72.37%**  **NPV=98%** |
| ≥ 6 | 23.81% | 97.38% | 21.88% | 96.49% | 27.66% | 96.90% | 11.32% | 96.33% | 31.44% | 93.73% | 69.31% | 83.41% |
| ≥ 7 | 14.29% | 98.79% | 7.81% | 98.30% | 17.02% | 98.66% | 7.55% | 98.17% | 23.71% | 95.96% | 52.91% | 90.66% |
| ≥ 8 | 9.52% | 99.52% | 4.69% | 99.26% | 10.64% | 99.25% | 5.66% | 98.79% | 17.53% | 97.46% | 34.92% | 95.48% |
| ≥ 9 | 2.38% | 99.90% | 3.13% | 99.61% | 2.13% | 99.68% | 3.77% | 99.41% | 12.89% | 98.58% | 18.78% | 98.21% |
| ≥ 10 | 0.00% | 99.99% | 1.56% | 99.87% | 2.13% | 99.92% | 3.77% | 99.74% | 8.25% | 99.57% | 9.26% | 99.49% |
| Note: PPV=Positive predictive values. NPV = Negative predictive values.  Sensitivity and specificity estimates of the SDQ emotional subscale are based on assessments at the concurrent age of the depression diagnoses (although note there is a slight age gap between SDQ and diagnosis assessments). All but the SDQ assessment at 25 years are based on parent-reports. Depression diagnoses at ages 7, 10, and 13 years are based on parent-reports, while diagnoses at 15 and 25 years are based on self-reports. Scores on the SDQ emotional subscale of 5 and above have been suggested to capture those with ‘high’ problems (see sdqinfo.org). Estimates in bold represent the optimum cut-point according to the balance of sensitivity and specificity. | | | | | | | | | | | | |

| **Supplementary Table 6: Sensitivity and specificity of the emotional subscale cutoff-points across development compared against Generalised Anxiety Disorder Diagnoses** | | | | | | | | |
| --- | --- | --- | --- | --- | --- | --- | --- | --- |
| Cut-point | Generalised Anxiety Disorder at 7 years | | Generalised Anxiety Disorder at 10 years | | Generalised Anxiety Disorder at 13 years | | Generalised Anxiety Disorder at 15/16 years | |
|  | Sensitivity | Specificity | Sensitivity | Specificity | Sensitivity | Specificity | Sensitivity | Specificity |
| ≥ 1 | 100% | 36.14% | 96.67% | 37.59% | 100.00% | 39.49% | 100.00% | 41.68% |
| ≥ 2 | 92.31% | 61.55% | 83.33% | 62.11% | 96.00% | 65.02% | **81.82%**  **PPV=1%** | **65.01%**  **NPV=>99%** |
| ≥ 3 | 84.62% | 77.60% | 80.00% | 77.36% | 96.00% | 79.78% | 68.18% | 78.44% |
| ≥ 4 | 76.92% | 87.61% | **76.67%**  **PPV=3%** | **87.11%**  **NPV=>99%** | **88.00%**  **PPV=3%** | **88.45%**  **NPV=>99%** | 40.91% | 87.16% |
| ≥ 5 | **76.92%**  **PPV=2%** | **93.73%**  **NPV=>99%** | 70.00%  PPV=4% | 93.19%  NPV=>99% | 60.00%  PPV=4% | 94.05%  NPV=>99% | 18.18%  PPV=1% | 92.46%  NPV=>99% |
| ≥ 6 | 46.15% | 97.34% | 53.33% | 96.54% | 48.00% | 96.95% | 13.64% | 96.28% |
| ≥ 7 | 38.46% | 98.79% | 30.00% | 98.35% | 28.00% | 98.66% | 13.64% | 98.15% |
| ≥ 8 | 30.77% | 99.53% | 20.00% | 99.28% | 16.00% | 99.24% | 13.64% | 98.80% |
| ≥ 9 | 0.00% | 99.89% | 10.00% | 99.62% | 8.00% | 99.70% | 9.09% | 99.41% |
| ≥ 10 | 0.00% | 99.99% | 6.67% | 99.88% | 4.00% | 99.92% | 4.55% | 99.72% |
| Note: PPV=Positive predictive values. NPV = Negative predictive values.  Sensitivity and specificity estimates of the SDQ emotional subscale are based on assessments at the concurrent age of the GAD diagnoses (although note there is a slight age gap between SDQ and diagnosis assessments). All but the SDQ assessment at 25 years are based on parent-reports. GAD diagnoses at ages 7, 10, and 13 years are based on parent-reports, while diagnoses at 15 years are based on self-reports. Scores on the SDQ emotional subscale of 5 and above have been suggested to capture those with ‘high’ problems (see sdqinfo.org). | | | | | | | | |

| **Supplementary Table 7: Sensitivity and specificity of the emotional subscale cutoff-points across development compared against any anxiety disorder diagnoses** | | | | | | | | |
| --- | --- | --- | --- | --- | --- | --- | --- | --- |
| Cut-point | Any Anxiety Disorder at 7 years | | Any Anxiety Disorder at 10 years | | Any Anxiety Disorder at 13 years | | Any Anxiety Disorder at 15/16 years | |
|  | Sensitivity | Specificity | Sensitivity | Specificity | Sensitivity | Specificity | Sensitivity | Specificity |
| ≥1 | 94.69% | 36.50% | 91.49% | 37.63% | 94.12% | 39.12% | 88.89% | 41.88% |
| ≥ 2 | 82.30% | 62.14% | 75.89% | 62.28% | 82.35% | 64.64% | **71.43%**  **PPV=3%** | **65.14%**  **NPV=>99%** |
| ≥ 3 | **68.14%**  **PPV=5%** | **78.17%**  **NPV=99%** | 64.54% | 77.57% | 70.59% | 79.56% | 50.79% | 78.52% |
| ≥ 4 | 57.52% | 88.18% | **56.74%**  **PPV=9%** | **87.49%**  **NPV=>99%** | **64.71%**  **PPV=8%** | **88.57%**  **NPV=>99%** | 38.10% | 87.42% |
| ≥ 5 | 44.25% | 94.21% | 48.23% | 93.68% | 49.41% | 94.25% | 26.98% | 92.68% |
| ≥ 6 | 26.55% | 97.63% | 32.62% | 96.85% | 30.59% | 97.08% | 20.63% | 96.47% |
| ≥ 7 | 16.81% | 98.98% | 17.73% | 98.54% | 18.82% | 98.84% | 14.29% | 98.22% |
| ≥ 8 | 8.85% | 99.62% | 10.64% | 99.40% | 11.76% | 99.38% | 9.52% | 98.84% |
| ≥ 9 | 1.77% | 99.91% | 6.38% | 99.69% | 3.53% | 99.75% | 4.76% | 99.43% |
| ≥ 10 | 0.00% | 99.99% | 2.13% | 99.89% | 1.18% | 99.93% | 3.17% | 99.74% |
| Note: PPV=Positive predictive values. NPV = Negative predictive values.  Sensitivity and specificity estimates of the SDQ emotional subscale are based on assessments at the concurrent age of the anxiety diagnosis (at ages 7, 10, 13, 15/16, and 25 years – although note there is a slight age gap between SDQ and diagnosis assessments). All but the SDQ assessment at 25 years are based on parent-reports. Any anxiety diagnosis at ages 7, 10 and 13 years are based on parent-reports, while diagnoses at 15 years are based on self-reports. Scores on the SDQ emotional subscale of 5 and above have been suggested to capture those with ‘high’ problems (see sdqinfo.org). | | | | | | | | |

| **Supplementary Table 8: Accuracy of identifying those meeting diagnostic criteria by optimal SDQ subscale cut-point** | | | | | | | | |
| --- | --- | --- | --- | --- | --- | --- | --- | --- |
| **Major Depressive Disorder** | | | **Generalised anxiety disorder** | | | **Any anxiety disorder** | | |
| Optimal cut-point | Met diagnostic criteria | Did not meet diagnostic criteria | Optimal cut-point | Met diagnostic criteria | Did not meet diagnostic criteria | Optimal cut-point | Met diagnostic criteria | Did not meet diagnostic criteria |
| **7 years** |  |  | **7 years** |  |  | **7 years** |  |  |
| (cut-off ≥ 3) | 26 (62%) | 1,578 (29%) | (cut-off ≥ 5) | 10 (77%) | 452 (6%) | (cut-off ≥ 3) | 78 (68%) | 1,541 (22%) |
| (cut-off < 3) | 16 (38%) | 5,509 (71%) | (cut-off < 5) | 3 (23%) | 6,753 (94%) | (cut-off < 3) | 36 (32%) | 5,517 (78%) |
| **10 years** |  |  | **10 years** |  |  | **10 years** |  |  |
| (cut-off ≥ 2) | 52 (81%) | 2,531 (38%) | (cut-off ≥ 4) | 23 (77%) | 882 (13%) | (cut-off ≥ 4) | 82 (57%) | 807 (13%) |
| (cut-off < 2) | 12 (19%) | 4,186 (62%) | (cut-off < 4) | 7 (23%) | 5,961 (87%) | (cut-off < 4) | 62 (43%) | 5,645 (87%) |
| **13 years** |  |  | **13 years** |  |  | **13 years** |  |  |
| (cut-off ≥ 3) | 37 (79%) | 1,197 (20%) | (cut-off ≥ 4) | 22 (88%) | 701 (12%) | (cut-off ≥ 4) | 56 (64%) | 630 (11%) |
| (cut-off < 3) | 10 (21%) | 4,773 (80%) | (cut-off < 4) | 3 (12%) | 5,366 (88%) | (cut-off < 4) | 31 (36%) | 4,884 (89%) |
| **15/16 years** |  |  | **15/16 years** |  |  | **15/16 years** |  |  |
| (cut-off ≥ 3) | 28 (52%) | 841 (22%) | (cut-off ≥ 2) | 18 (82%) | 1,381 (35%) | (cut-off ≥ 2) | 46 (72%) | 1,352 (35%) |
| (cut-off < 3) | 26 (48%) | 3,054 (78%) | (cut-off < 2) | 4 (18%) | 2,541 (65%) | (cut-off < 2) | 18 (28%) | 2,526 (65%) |
| **25 years** |  |  | - |  |  | - |  |  |
| (cut-off ≥ 3) | 115 (59%) | 627 (27%) | - |  |  | - |  |  |
| (cut-off < 3) | 79 (41%) | 1,700 (73%) | - |  |  | - |  |  |
| **25 years (self)** |  |  | - |  |  | - |  |  |
| (cut-off ≥ 5) | 314 (83%) | 1,021 (28%) | - |  |  | - |  |  |
| (cut-off < 5) | 64 (17%) | 2,674 (72%) | - |  |  | - |  |  |
| Note: SDQ assessments are based on the concurrent age of the diagnosis, however there is some gap between assessments. All SDQ assessments are based on parent-reports unless stated otherwise. Diagnoses at ages 7, 10 and 13 years are based on parent-reports, while diagnoses at 15 and 25 years are based on self-reports. | | | | | | | | |

| **Supplementary Table 9: Discrimination of those with versus without DAWBA diagnoses for the depressive item by sex** | | | | | | | | | | | | |
| --- | --- | --- | --- | --- | --- | --- | --- | --- | --- | --- | --- | --- |
|  | **Major Depressive Disorder** | | | **Generalised Anxiety Disorder** | | | **Any anxiety disorder** | | | **Attention Deficit Hyperactivity Disorder (ADHD) or any behavioural disorder** | | |
| **Age** | **Males** | **Females** | **Diff** | **Males** | **Females** | **Diff** | **Males** | **Females** | **Diff** | **Males** | **Females** | **Diff** |
|  | AUC  (95% CI) | AUC  (95% CI) | χ^2^_(1)_, p-value | AUC  (95% CI) | AUC  (95% CI) | χ^2^_(1)_, p-value | AUC  (95% CI) | AUC  (95% CI) | χ^2^_(1)_, p-value | AUC  (95% CI) | AUC  (95% CI) | χ^2^_(1)_, p-value |
| 7 years | 0.76  (0.66, 0.86) | 0.61  (0.48, 0.73) | 3.71,  **<0.05** | 0.80  (0.64, 0.96) | 0.75  (0.43, 1.00) | 0.08,  0.77 | 0.66  (0.59, 0.72) | 0.59  (0.52, 0.66) | 1.89,  0.17 | 0.60  (0.57, 0.64) | 0.62  (0.56, 0.69) | 0.35,  0.55 |
| 10 years | 0.67  (0.58, 0.76) | 0.65  (0.56, 0.75) | 0.08,  0.78 | 0.78  (0.66, 0.90) | 0.71  (0.54, 0.88) | 0.36,  0.55 | 0.70  (0.64, 0.76) | 0.64  (0.58, 0.70) | 2.05,  0.15 | 0.62  (0.58, 0.66) | 0.66  (0.59, 0.72) | 0.75,  0.39 |
| 13 years | 0.75  (0.65, 0.85) | 0.76  (0.66, 0.86) | 0.00,  0.95 | 0.76  (0.63, 0.90) | 0.88  (0.78, 0.97) | 1.91,  0.17 | 0.70  (0.63, 0.79) | 0.68  (0.60, 0.75) | 0.31,  0.58 | 0.60  (0.55, 0.64) | 0.62  (0.57, 0.68) | 0.49,  0.49 |
| 15/16 years | 0.55  (0.45, 0.66) | 0.62  (0.53, 0.70) | 0.86,  0.35 | 0.45  (0.45, 0.46) | 0.75  (0.65, 0.86) | 29.66,  **<0.001** | 0.45  (0.45, 0.46) | 0.67  (0.60, 0.74) | 35.99  **<0.001** | 0.60  (0.54, 0.66) | 0.71  (0.64, 0.78) | 5.35,  **<0.05** |
| 25 years | 0.68  (0.60, 0.76) | 0.68  (0.64, 0.72) | 0.00,  0.97 | - | - | - | - | - | - | - | - | - |
| 25 years  (self) | 0.89  (0.85, 0.92) | 0.85  (0.83, 0.87) | 3.20,  0.07 | - | - | - | - | - | - | - | - | - |
| Note: *Any behavioural disorder includes Conduct Disorder (CD) and Oppositional Defiant Disorder (ODD). All DAWBA diagnoses at ages 7, 10 and 13 years are based on parent-reports, while diagnoses at 15 and 25 are based on self-reports. | | | | | | | | | | | | |

| **Supplementary Table 10: Discrimination of those with versus without DAWBA diagnoses for the worry item by sex** | | | | | | | | | | | | |
| --- | --- | --- | --- | --- | --- | --- | --- | --- | --- | --- | --- | --- |
|  | **Major Depressive Disorder** | | | **Generalised Anxiety Disorder** | | | **Any anxiety disorder** | | | **Attention Deficit Hyperactivity Disorder (ADHD) or any** **behavioural disorder*** | | |
| **Age** | **Males** | **Females** | **Diff** | **Males** | **Females** | **Diff** | **Males** | **Females** | **Diff** | **Males** | **Females** | **Diff** |
|  | AUC  (95% CI) | AUC  (95% CI) | χ^2^_(1)_, p-value | AUC  (95% CI) | AUC  (95% CI) | χ^2^_(1)_, p-value | AUC  (95% CI) | AUC  (95% CI) | χ^2^_(1)_, p-value | AUC  (95% CI) | AUC  (95% CI) | χ^2^_(1)_, p-value |
| 7 years | 0.77  (0.67, 0.86) | 0.67  (0.53, 0.80) | 1.35,  0.25 | 0.80  (0.65, 0.94) | 0.94  (0.86, 1.00) | 2.93,  0.09 | 0.71  (0.64, 0.77) | 0.71  (0.64, 0.79) | 0.01, 0.94 | 0.57  (0.53, 0.60) | 0.59  (0.53, 0.65) | 0.45,  0.50 |
| 10 years | 0.75  (0.67, 0.83) | 0.75  (0.67, 0.84) | 0.01,  0.93 | 0.83  (0.74, 0.93) | 0.79  (0.63, 0.96) | 0.19,  0.67 | 0.75  (0.69, 0.81) | 0.66  (0.60, 0.73) | 3.40,  0.07 | 0.61  (0.57, 0.66) | 0.61  (0.55, 0.68) | 0.00,  0.98 |
| 13 years | 0.70  (0.60, 0.81) | 0.77  (0.67, 0.88) | 0.91,  0.34 | 0.85  (0.75, 0.96) | 0.89  (0.85, 0.92) | 0.32,  0.57 | 0.76  (0.68, 0.83) | 0.74  (0.66, 0.81) | 0.10,  0.75 | 0.60  (0.56, 0.65) | 0.63  (0.57, 0.69) | 0.34,  0.56 |
| 15/16 years | 0.70  (0.56, 0.84) | 0.63  (0.54, 0.72) | 0.81,  0.37 | 0.85  (0.83, 0.86) | 0.68  (0.56, 0.80) | 7.70,  **<0.01** | 0.51  (0.36, 0.66) | 0.64  (0.56, 0.71) | 2.38, 0.12 | 0.63  (0.56, 0.69) | 0.66  (0.58, 0.73) | 0.48,  0.49 |
| 25 years (parent) | 0.64  (0.56, 0.72) | 0.67  (0.62, 0.71) | 0.32, 0.57 | - | - | - | - | - | - | - | - | - |
| 25 years (self) | 0.77  (0.73, 0.82) | 0.71  (0.69, 0.74) | 5.27,  **<0.05** | - | - | - | - | - | - | - | - | - |
| Note: *Any behavioural disorder includes Conduct Disorder (CD) and Oppositional Defiant Disorder (ODD). All DAWBA diagnoses at ages 7, 10 and 13 years are based on parent-reports, while diagnoses at 15 and 25 are based on self-reports. | | | | | | | | | | | | |

| **Supplementary Table 11: Comorbidity of DAWBA Depressive and Anxiety disorders by sex** | | | | | | | | | | |
| --- | --- | --- | --- | --- | --- | --- | --- | --- | --- | --- |
| Age | **Major Depressive disorder** | | | | **Generalised Anxiety Disorder** | | | **Any anxiety disorder** | | |
|  | N | With disorder (%) | Diagnosed and have GAD (%) | Diagnosed and have any anxiety disorder (%) | N | With disorder (%) | Diagnosed and have MDD (%) | N | With disorder (%) | Diagnosed and have MDD (%) |
| **All** |  |  |  |  |  |  |  |  |  |  |
| 7 years | 7,987 | 52 (0.7%) | 8 (15.4%) | 18 (34.6%) | 8,098 | 17 (0.2%) | 8 (47.1%) | 8,041 | 138 (1.7%) | 17 (13.0%) |
| 10 years | 7,560 | 74 (1.0%) | 12 (16.2%) | 21 (28.4%) | 7,674 | 33 (0.4%) | 12 (36.4%) | 8,063 | 160 (2.2%) | 21 (13.0%) |
| 13 years | 6,871 | 58 (0.8%) | 12 (20.7%) | 19 (32.8%) | 6,969 | 31 (0.4%) | 12 (38.7%) | 6,401 | 104 (1.6%) | 19 (18.3%) |
| 15 years | 5,293 | 86 (1.6%) | 13 (15.1%) | 22 (25.6%) | 5,289 | 38 (0.7%) | 13 (34.2%) | 5,275 | 101 (1.9%) | 22 (21.8%) |
| **Males** |  |  |  |  |  |  |  |  |  |  |
| 7 years | 4,090 | 30 (0.7%) | 6 (20.0%) | 10 (33.3%) | 4,158 | 14 (0.3%) | 6 (42.9%) | 4,130 | 82 (2.0%) | 10 (12.2%) |
| 10 years | 3,804 | 41 (1.1%) | 2 (4.9%) | 14 (34.1%) | 3,869 | 21 (0.5%) | 8 (38.1%) | 3,691 | 82 (2.2%) | 14 (17.1%) |
| 13 years | 3,429 | 31 (0.9%) | 6 (19.4%) | 10 (32.3%) | 3,497 | 14 (0.4%) | 6 (42.9%) | 3,185 | 48 (1.5%) | 10 (20.0%) |
| 15 years | 2,500 | 24 (1.0%) | 1 (4.2%) | 2 (8.3%) | 2,499 | 4 (0.2%) | 1 (25%) | 2,496 | 17 (0.7%) | 2 (11.8%) |
| **Females** |  |  |  |  |  |  |  |  |  |  |
| 7 years | 3,897 | 22 (0.6%) | 2 (9.0%) | 8 (36.4%) | 3,940 | 3 (0.08%) | 2 (66.6%) | 3,911 | 56 (1.4%) | 10 (17.9%) |
| 10 years | 3,756 | 33 (0.9%) | 2 (6.1%) | 7 (21.2%) | 3,805 | 12 (0.3%) | 4 (33.3%) | 3,662 | 78 (2.1%) | 7 (8.9%) |
| 13 years | 3,442 | 27 (0.8%) | 6 (22.2%) | 9 (33.3%) | 3,472 | 17 (0.5%) | 6 (35.3%) | 3,216 | 56 (1.5%) | 9 (19.6%) |
| 15 years | 2,785 | 62 (2.2%) | 12 (19.4%) | 20 (32.3%) | 2,782 | 34 (1.2%) | 12 (35.3%) | 2,779 | 84 (3.0%) | 20 (23.8%) |

**Supplementary Figure 1:** ROC analyses for emotional subscale predicting any anxiety diagnosis across development

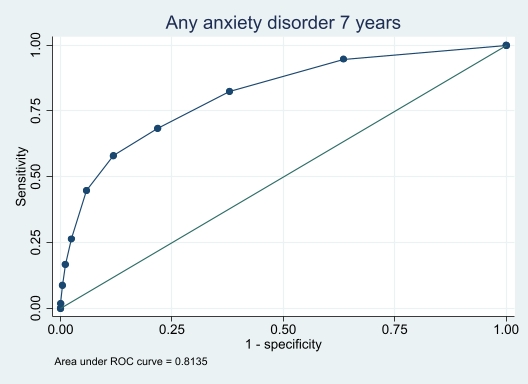

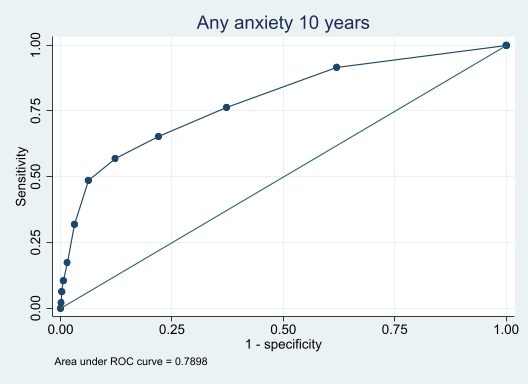

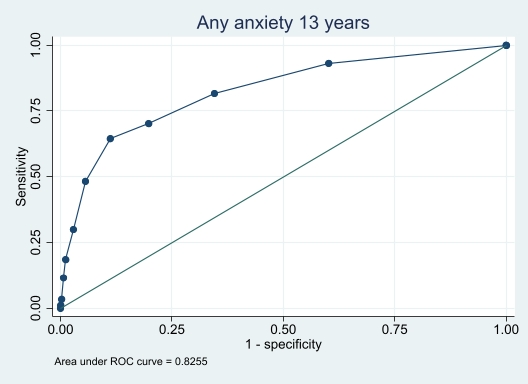

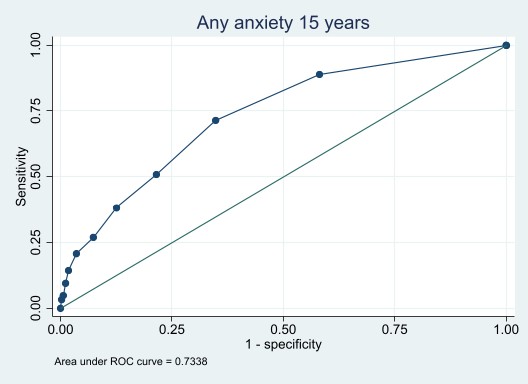

**Supplementary Figure 2:** ROC analyses for emotional subscale predicting Attention Deficit Hyperactivity Disorder or any behavioural diagnosis across development

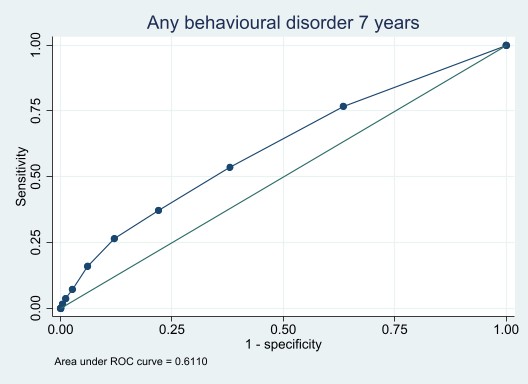

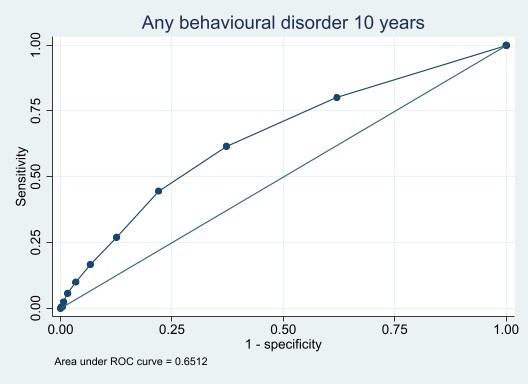

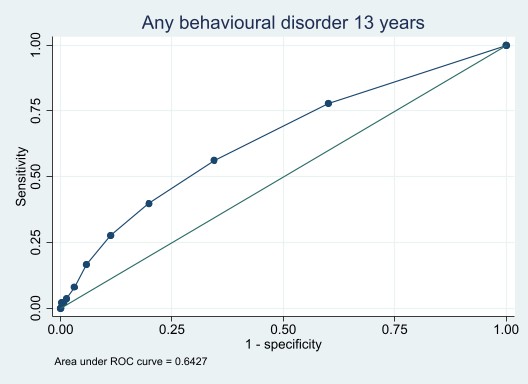

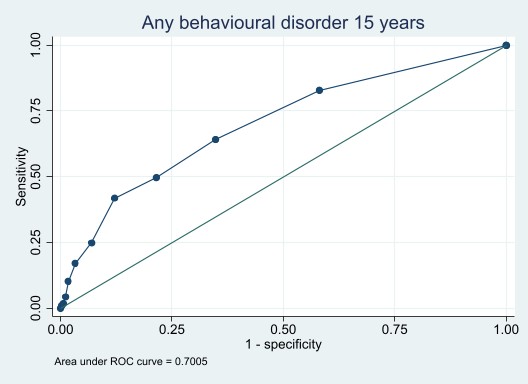
